# The HIV/STI Epidemic Potential of Dynamic Sexual Networks of Men Who Have Sex With Men in Atlanta and San Francisco

**DOI:** 10.1101/2020.10.12.20211540

**Authors:** Emeli J. Anderson, Kevin M. Weiss, Martina M. Morris, Travis H. Sanchez, Pragati Prasad, Samuel M. Jenness

## Abstract

**Background:** The potential speed through which a pathogen may circulate in a network is a function of network connectivity. Network features like degree (number of ongoing partnerships) determine the cross-sectional network connectivity. The overall transmission potential of a pathogen involves connectivity over time, which can be measured using the forward reachable path (FRP). We modeled dynamic sexual networks of MSM in San Francisco and Atlanta to estimate the FRP as a predictor of HIV/STI epidemic potential.

**Methods:** We used exponential random graph models to obtain parameter estimates for each city’s sexual network and then simulated the complete networks over time. The FRP was estimated in each city overall and stratified by demographics.

**Results:** The overall mean and median FRPs were higher in San Francisco than in Atlanta, suggesting a greater epidemic potential for HIV and STIs in San Francisco. At one year, in both cities, the average FRP among casual partnerships was highest in the youngest age group and lowest in the oldest age group, contrasting with the cross-sectional network parameters we estimated, where the youngest age category had the lowest mean degree and the oldest age category had the highest mean degree.

**Conclusions:** The FRP results correspond to the observed STI epidemics but not HIV epidemics between the cities. In San Francisco, rates of HIV have been declining over the last few years, whereas they have been steady in Atlanta. The FRP by age group resulted in fundamentally different conclusions about connectivity in the network compared with the cross-sectional network measures.

## INTRODUCTION

HIV and other sexually transmitted infections (STIs) disproportionately affect men who have sex with men (MSM). In 2018, 69% of all new HIV diagnoses and 64% of reported primary and secondary syphilis infections occurred among MSM, despite representing less than 5% of the total U.S. population.^1–3^ Biomedical HIV prevention tools, including HIV preexposure prophylaxis (PrEP), have scaled-up in recent years while treatment for bacterial STIs has been available for decades (i.e., antibiotics).^4,5^ However, disparities in both epidemics persist by age, race, and geography.^5–9^ Studying dynamic sexual networks, graphs that represent the sexual connections between people in a population over time, can provide insight into the transmission dynamics that result in the continued HIV and STI disparities in MSM.

Individual-level risk factors, such as condomless anal intercourse, have been insufficient to explain the high HIV/STI burden among MSM.^8,10,11^ The framework of sexual networks has furthered our understanding of the potential drivers of HIV and STIs at the population level.^9,12,13^ Network connectivity, which is a determinant of the epidemic potential of an STI, is a function of cross-sectional network features like degree (number of ongoing partnerships at a point in time).^14^ Partnership concurrency (degree ≥ 2) facilitates HIV/STI transmission, particularly during periods of heightened biological transmissibility, such as during the acute phase of HIV infection.^12,15,16^ Conversely, networks with high levels of assortative mixing, in which individuals choose partners with attributes similar to themselves, may slow transmission by restricting an epidemic within a smaller, high prevalence group.^11,22^ Estimating cross-sectional network features can indicate the potential transmission risk within the network. However, this represents a simplification of the actual temporal coevolution of networks and pathogen spread.^19–22^ The overall effect of cross-sectional network features on the transmission potential of HIV/STIs in a sexual network depends on their interaction with the duration of partnerships, which is only observed in dynamic networks.^15^

Within sexual networks, the *forward reachable path* (FRP) is one potential informative temporal network measure.^23,24^ The FRP quantifies the maximum number of men each man is connected to directly, through his own partners, and indirectly, through partners of partners over time. The FRP can be thought of as an upper threshold for epidemic spread, representing how far an epidemic would spread if the transmission probability per contact were 100%. Although this is not realistic for projecting future STI impact, the FRP across groups (like age, race/ethnicity, and geography) may provide a useful measure of epidemic potential that is estimated using a data-driven approach, without the assumptions of a full-scale infectious disease transmission model.^25^

The FRP as a measure in temporal sexual networks is relatively recent due to advances in statistical methodology allowing for estimation of the whole network of a population over time.^24^ The importance of the temporal interactions between network features like degree, concurrency, and partnership duration has been shown in dynamic network models using simulated data. Generally, the lower the mean degree in the population, the less connected the network, both cross-sectionally and temporally. However, in networks with a low mean degree, the more skewed the distribution of individual degrees, the greater the temporal reachability within the network.^14,26–28^ Further, the interaction between cross-sectional degree and partnership duration results in emergent properties in the temporal network relating to the epidemic threshold (i.e., the minimum level of connectivity required for an epidemic to occur) and the growth rate of the FRP.^23^ In particular, networks with concurrency result in a lower epidemic threshold and a higher growth rate of the FRP compared to networks of just monogamous partnerships. This result supports observations from an earlier study using empirical data, which found that going from a network with only monogamous partnerships to one in which concurrency was set to observed levels could double the FRP.^12^ These studies show how network structure critically determines temporal network connectivity. One question is whether the FRP might be able to predict relative differences in the observed HIV and STI epidemics in stratifications of interest, such as geography.

Here, we examine the FRP in sexual network models using empirical data from San Francisco and Atlanta to identify whether the epidemic potential across geographic locations might be predictive of the relative HIV and STI epidemics in those settings. The HIV and STI epidemiology in these cities provides insight into contrasting disease trends and public health responses.^29^ Overall HIV rates declined in San Francisco by 13% between 2017 and 2018. ^30^ In Georgia, HIV rates have remained stable over the last decade and Atlanta currently has the second highest rate of new HIV diagnoses among major U.S. cities.^29,31^ Conversely to the HIV trends, the rate of gonorrhea infection among men in San Francisco was almost double the rate among men in Atlanta in 2018 and the rate has been increasing faster in San Francisco than in Atlanta.^1^ The rate of primary and secondary syphilis infection among men has remained mostly stable in both cities, though in 2018 it was slightly higher in San Francisco. Given these differences, examining the FRP in each city’s network may provide insight into the interaction between network epidemic potential and the public health interventions implemented in each city.

For this study, we modeled dynamic sexual networks of MSM in San Francisco and Atlanta to estimate the FRP as a predictor of epidemic potential in each city. We sought to quantify how the FRP evolved over time, both overall and stratified by age and race/ethnicity. Our goal was to understand the contribution of dynamic sexual network structure to the evolving HIV/STI transmission dynamics in these two cities.

## METHODS

### Study Design

We used network data from ARTnet, a web-based study of MSM conducted between 2017 and 2019.^32^ Participants for ARTnet were recruited through the American Men’s Internet Survey (AMIS), an annual web-based national survey.^33^ ARTnet eligibility criteria included male sex at birth, a current male gender identity, lifetime history of sexual activity with another man, and age between 15 and 65. Participants provided information on their sexual partnerships, but the partners were not sampled directly. After deduplication within and across survey waves, ARTnet had a final sample size of 4,904 participants reporting on 16,198 sexual partnerships. The study was approved by the Emory University Institutional Review Board.

To estimate the FRPs in each city’s network, we followed a 4-step procedure summarized in **Figure 1** and described below. The procedure is data-driven using empirical data from a sample to make inferences about the complete dynamic network of a target population. This process contrasts with transmission-based infectious disease models, in which estimated parameters are used to generate simulated data. A detailed description of these methods can be found in Krivtisky and Morris 2017.^20^ Note that we do not directly model any infectious process but instead assume that every node in the path would become infected (i.e. a 100% transmission probability). This provides an upper limit on the size of a potential epidemic in the network. Analyses were conducted using the Statnet suite of R packages.^34^ Reproducible code may be found at: https://github.com/EpiModel/NetAnalysis-SF-ATL.

**Figure 1.**
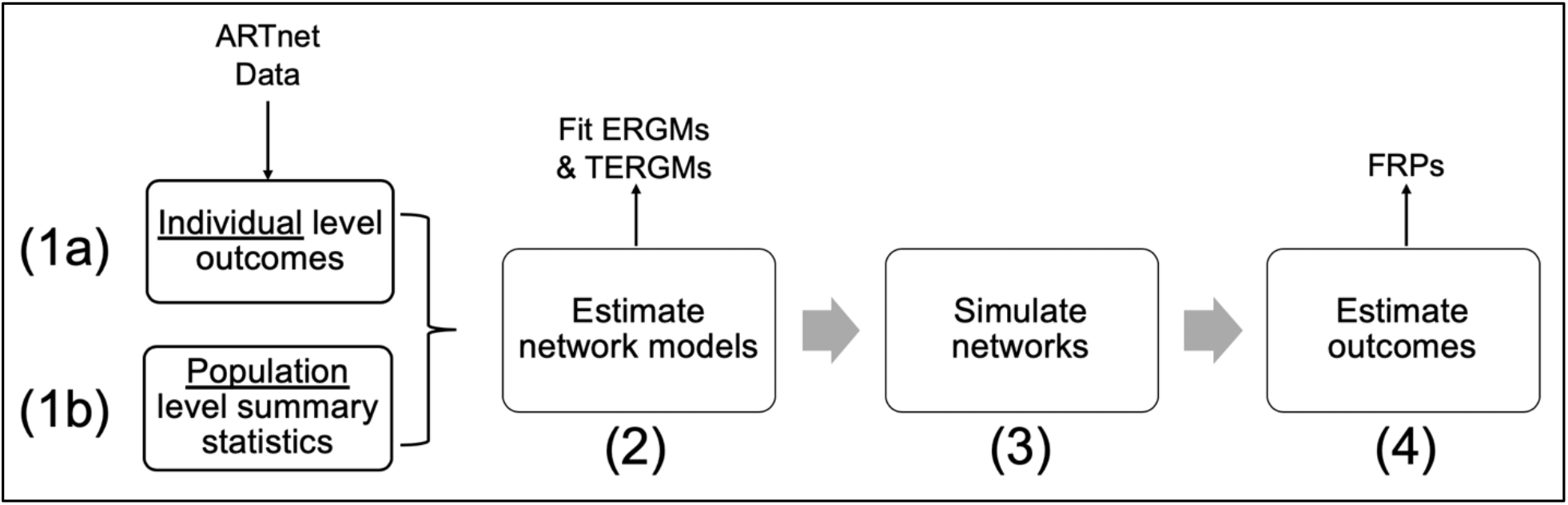
The network analysis process. Step (1a) Estimate individual-level network statistics. Step (1b) Combine estimates from 1a with population-level demographic weights to estimate population-level summary statistics. Step (2) Fit network models using exponential random graph models (ERGMs) for one-time partnerships and temporal ERGMs for partnerships with duration (main and casual partnerships). Inputs for these models come from step 1b. Step (3) Simulate the complete networks over time for each city. Step (4) estimate and compare the forward reachable paths (FRPs) for each city.

### Step 1a: Individual-Level Network Measures

The individual-level network measures are provided in **Table 1**. Descriptive statistics of the participants and their reported partnerships are described elsewhere.^32^ Partnerships were categorized as main (partner considered a boyfriend, significant other, or life partner), casual (an ongoing relationship, but not a main partner), and one-time contacts. Age was categorized into 10-year intervals, ranging from 15 to 64. Race/ethnicity was dichotomized as Black non-Hispanic/Hispanic and White non-Hispanic/Other. Cross-partnership degree for ongoing partnerships (main and casual) represented the number of partnerships in each partnership category as a function of the number in the other type.

**Table 1.** Network Parameters for Sexual Networks of Men Who Have Sex with Men (MSM) in San Francisco and Atlanta.

| Parameter | Main Partnerships |  | Casual Partnerships |  | One-Time Partnerships |  |
| --- | --- | --- | --- | --- | --- | --- |
|  | San Francisco | Atlanta | San Francisco | Atlanta | San Francisco | Atlanta |
| **Mean Degree** |  |  |  |  |  |  |
| *Overall* | 0.47 | 0.40 | 0.88 | 0.54 | 0.19 | 0.08 |
| *By Age Group* |  |  |  |  |  |  |
| 15–24 | 0.45 | 0.37 | 0.46 | 0.30 | 0.12 | 0.05 |
| 25–34 | 0.57 | 0.47 | 0.75 | 0.48 | 0.19 | 0.08 |
| 35–44 | 0.54 | 0.45 | 0.96 | 0.61 | 0.22 | 0.09 |
| 45–54 | 0.45 | 0.37 | 1.09 | 0.70 | 0.23 | 0.09 |
| 55–64 | 0.34 | 0.28 | 1.16 | 0.74 | 0.21 | 0.09 |
| *By Race* |  |  |  |  |  |  |
| Black/Hispanic | 0.44 | 0.37 | 0.89 | 0.55 | 0.17 | 0.07 |
| White/Other | 0.48 | 0.40 | 0.88 | 0.54 | 0.20 | 0.08 |
| *By Cross Partnership Degree* |  |  |  |  |  |  |
| 0 | 0.49 | 0.63 | 0.55 | 0.61 | - | - |
| 1 | 0.25 | 0.22 | 0.43 | 0.38 | - | - |
| 2 | 0.15 | 0.11 | 0.02 | 0.10 | - | - |
| 3 | 0.11 | 0.03 | - | - | - | - |
| **Assortative Age Mixing** |  |  |  |  |  |  |
| 15–24 | 0.77 | 0.80 | 0.60 | 0.57 | 0.55 | 0.56 |
| 25–34 | 0.67 | 0.70 | 0.48 | 0.44 | 0.43 | 0.45 |
| 35–44 | 0.54 | 0.58 | 0.36 | 0.32 | 0.32 | 0.33 |
| 45–54 | 0.41 | 0.44 | 0.25 | 0.22 | 0.23 | 0.24 |
| 55–64 | 0.29 | 0.32 | 0.17 | 0.15 | 0.16 | 0.16 |
| **Assortative Race Mixing** |  |  |  |  |  |  |
| Black/Hispanic | 0.47 | 0.55 | 0.52 | 0.51 | 0.53 | 0.54 |
| White/Other | 0.78 | 0.83 | 0.71 | 0.70 | 0.75 | 0.76 |
| **One-Time Partner Rates by Risk Quintile** |  |  |  |  |  |  |
| 20% | - | - | - | - | 0.00 | 0.00 |
| 40% | - | - | - | - | 0.00 | 0.00 |
| 60% | - | - | - | - | 0.03 | 0.01 |
| 80% | - | - | - | - | 0.11 | 0.04 |
| 100% | - | - | - | - | 0.83 | 0.33 |
| **Mean Duration (Weeks)** |  |  |  |  |  |  |
| Different age partner | 216.9 | 219.3 | 100.9 | 104.6 | - | - |
| Same age partner: 15–24 | 70.7 | 71.5 | 46.7 | 48.3 | - | - |
| Same age partner: 25–34 | 246.9 | 249.8 | 67.3 | 69.7 | - | - |
| Same age partner: 35–44 | 520.9 | 526.8 | 106.1 | 110.0 | - | - |
| Same age partner: 45–54 | 664.4 | 672.0 | 149.7 | 155.2 | - | - |
| Same age partner: 55–64 | 1154.7 | 1167.9 | 139.9 | 145.0 | - | - |

The individual-level network measures included mean degree (overall and stratified by age, race/ethnicity, and cross partnership degree), one-time partnership acquisition rates, partnership durations, and preferential mixing by age and race. Mean degree was calculated as the average of the number of ongoing partnerships each individual reported. One-time partnerships were described by a weekly partnership acquisition rate calculated by dividing the total number of non-persistent past-year partners by 52. We further categorized rates of one-time partnership acquisition by a risk quintile to account for the right-skewed distribution of partnerships.^32^ Partnership durations (in weeks) for main and casual partnerships were estimated from partnership age (the difference between the survey date and the partnership start date) for extant partnerships. Partnership age has been shown to be an unbiased estimator of duration under statistical assumptions of exponentially distributed durations.^32^ Preferential mixing by age and race/ethnicity was quantified as the proportion of partnerships that were between individuals in the same demographic category.

### Step 1b: Population-Level Summary Statistics

To calculate population-level summary statistics we multiplied the individual-level measures from Step 1a by the race distribution within each city based on Census data and a population size of 10,000 MSM.^35^ Although this population size is smaller than the estimated MSM population size in each city (approximately 145,972 and 102,642 in San Francisco and Atlanta core-based statistical areas, respectively)^36^, the FRP measures are standardized so the choice of population size here was made primarily based on computational efficiency.

### Step 2: Network Estimation

Using the population-level summary statistics as data points, we fit ERGMs to estimate the generative properties of the sexual partnership networks. The estimand of an ERGM is the log odds of a partnership between each pair (or dyad) in the network, conditional on the rest of the network. A separate model was fit for each partnership type for each city. For one-time partnerships, cross-sectional ERGMs were fit. For persistent (main/casual) partnerships, temporal ERGMs (TERGMs) were fit. TERGMs model the likelihood of formation and dissolution of partnerships per unit time, whereas ERGMs model formation without partnership duration. Covariates for each model included nodal attributes (age, race/ethnicity, cross-partnership degree, and number with concurrent partnerships) and dyadic attributes (count of same-age and same-race partnerships).

### Step 3: Network Simulation

Parameters estimated from the ERGMs were then used to simulate complete dynamic networks in weekly time steps for five years. In the simulation, edges form and dissolve based on the model parameters. Nodal attributes for age and cross-partnership degree were updated each week. No exogenous population processes were implemented (e.g., mortality) so the total population size remained constant at 10,000.

### Step 4: Forward Reachable Path Estimation

The FRP is an individual measure that quantifies the size of the temporally ordered chain of edges for each person in the network. We demonstrate how it is calculated in the Supplemental Appendix [LINK]. The main outcome was the cumulative FRP, estimated weekly for each node. Analyses provide estimates of the mean, median, and interquartile range (IQR) for the distribution of the FRP measures across nodes in the network. We calculated FRPs for the combined partnership network, the sub-networks of each partnership type, and stratified by race/ethnicity and age. For the latter, we calculated the FRP for each node within the category of interest and estimated summary measures across the group.

## RESULTS

The individual-level network statistics are provided in **Table 1**. For main and casual partnerships, overall and within age and race/ethnicity categories, the mean degree was higher in San Francisco than in Atlanta (overall mean degree of 0.47 for main and 0.88 for casual in San Francisco and 0.40 for main and 0.54 for casual in Atlanta). Similarly, the one-time partnership formation rates were higher in San Francisco than in Atlanta (overall rates of 0.19 and 0.08, respectively).

For both Atlanta and San Francisco, the mean degrees and one-time partnership formation rates by race/ethnicity were not substantively different from the overall results, though there were some differences by age group. Among casual partnerships the oldest age group had the highest mean degree (1.16 for San Francisco and 0.74 for Atlanta) and the youngest age group had the lowest mean degree (0.46 for San Francisco and 0.30 for Atlanta). Age and race assortativity were similar between the cities. Assortativity decreased with age, where individuals in the youngest age group had the highest probability of being in a partnership with someone in the same age group and those in the oldest age group had the lowest probability. Within race/ethnicity categories, those in the White/Other group had a higher probability of a same-race partnership than those in the Black/Hispanic group.

The distribution of individual FRPs by partnership type and city can be found in **Figure 2**. Across partnership types, the FRPs among main partnerships rose slowly and reached few individuals (less than 30 in both cities). For San Francisco, the FRPs among casual partnerships rose more slowly than one-time partnership FRPs, but the final FRPs were higher among casual partnerships. In Atlanta, one-time partnerships reached more individuals and did so faster than casual partnerships. Figure 2 also shows the wide spread of individual FRPs, indicating a substantial degree of variability in the FRPs depending on which node started the path. This variability can also be seen in the IQR estimates in **Table 2**, which includes summary results of the FRPs at one year. For instance, for one-time partnerships, the IQR for the FRP at one year ranged from one to 4,514 for San Francisco and from one to 790 for Atlanta.

**Table 2.** The Forward Reachable Path at One Year by Partnership Type in San Francisco and Atlanta.

|  | Overall |  |  | Main |  |  | Casual |  |  | One-Time |  |  |
| --- | --- | --- | --- | --- | --- | --- | --- | --- | --- | --- | --- | --- |
|  | Mean | Median | IQR | Mean | Median | IQR | Mean | Median | IQR | Mean | Median | IQR |
| ***San Francisco*** |  |  |  |  |  |  |  |  |  |  |  |  |
| Overall | 8200 | 9121 | 8999 – 9121 | 1.7 | 2.0 | 1.0 – 2.0 | 145 | 12 | 1 – 134 | 2059 | 65 | 1 – 4514 |
| Age |  |  |  |  |  |  |  |  |  |  |  |  |
| 15–24 | 8432 | 9121 | 9028 – 9121 | 1.8 | 2.0 | 1.0 – 2.0 | 190 | 36 | 2 – 196 | 1835 | 1 | 1 – 4435 |
| 25–34 | 8229 | 9107 | 8999 – 9121 | 1.8 | 2.0 | 1.0 – 2.0 | 115 | 5 | 1 – 73 | 2010 | 4 | 1 – 4504 |
| 35–44 | 8367 | 9121 | 9064 – 9121 | 1.7 | 2.0 | 1.0 – 2.0 | 146 | 14 | 1 – 147 | 2155 | 869 | 1 – 4541 |
| 45–54 | 8433 | 9121 | 9064 – 9121 | 1.6 | 1.0 | 1.0 – 2.0 | 173 | 23 | 2 – 173 | 2257 | 2541 | 1 – 4576 |
| 55–64 | 7529 | 9088 | 8660 – 9121 | 1.4 | 1.0 | 1.0 – 2.0 | 101 | 3 | 1 – 61 | 2040 | 75 | 1 – 4505 |
| Race |  |  |  |  |  |  |  |  |  |  |  |  |
| Black/Hispanic | 8239 | 9121 | 9017 – 9121 | 1.6 | 2.0 | 1.0 – 2.0 | 157 | 18 | 1 – 157 | 2093 | 178 | 1 – 4507 |
| White/Other | 8190 | 9121 | 8999 – 9121 | 1.7 | 2.0 | 1.0 – 2.0 | 142 | 11 | 1 – 125 | 2051 | 46 | 1 – 4519 |
| ***Atlanta*** |  |  |  |  |  |  |  |  |  |  |  |  |
| Overall | 5159 | 7081 | 12 - 7448 | 1.5 | 1.0 | 1.0 – 2.0 | 8 | 2 | 1 – 8 | 505 | 1 | 1 – 790 |
| Age |  |  |  |  |  |  |  |  |  |  |  |  |
| 15–24 | 4889 | 6932 | 30 – 7448 | 1.6 | 1.0 | 1.0 – 2.0 | 10 | 2 | 1 – 11 | 283 | 1 | 1 – 15 |
| 25–34 | 5269 | 7080 | 420 – 7448 | 1.6 | 2.0 | 1.0 – 2.0 | 7 | 1 | 1 – 6 | 533 | 1 | 1 – 959 |
| 35–44 | 5580 | 7249 | 689 – 7448 | 1.6 | 2.0 | 1.0 – 2.0 | 9 | 2 | 1 – 9 | 562 | 1 | 1 – 1015 |
| 45–54 | 5452 | 7249 | 5 – 7448 | 1.5 | 1.0 | 1.0 – 2.0 | 10 | 2 | 1 – 11 | 619 | 1 | 1 – 1225 |
| 55–64 | 4599 | 6920 | 4 – 7448 | 1.3 | 1.0 | 1.0 – 2.0 | 6 | 1 | 1 – 4 | 526 | 1 | 1 – 901 |
| Race |  |  |  |  |  |  |  |  |  |  |  |  |
| Black/ Hispanic | 5085 | 7074 | 55 – 7448 | 1.5 | 1.0 | 1.0 – 2.0 | 8 | 2 | 1 – 8 | 492 | 1 | 1 – 718 |
| White/Other | 5258 | 7112 | 10 – 7448 | 1.5 | 1.0 | 1.0 – 2.0 | 8 | 1 | 1 – 8 | 522 | 1 | 1 – 864 |

**Figure 2.**
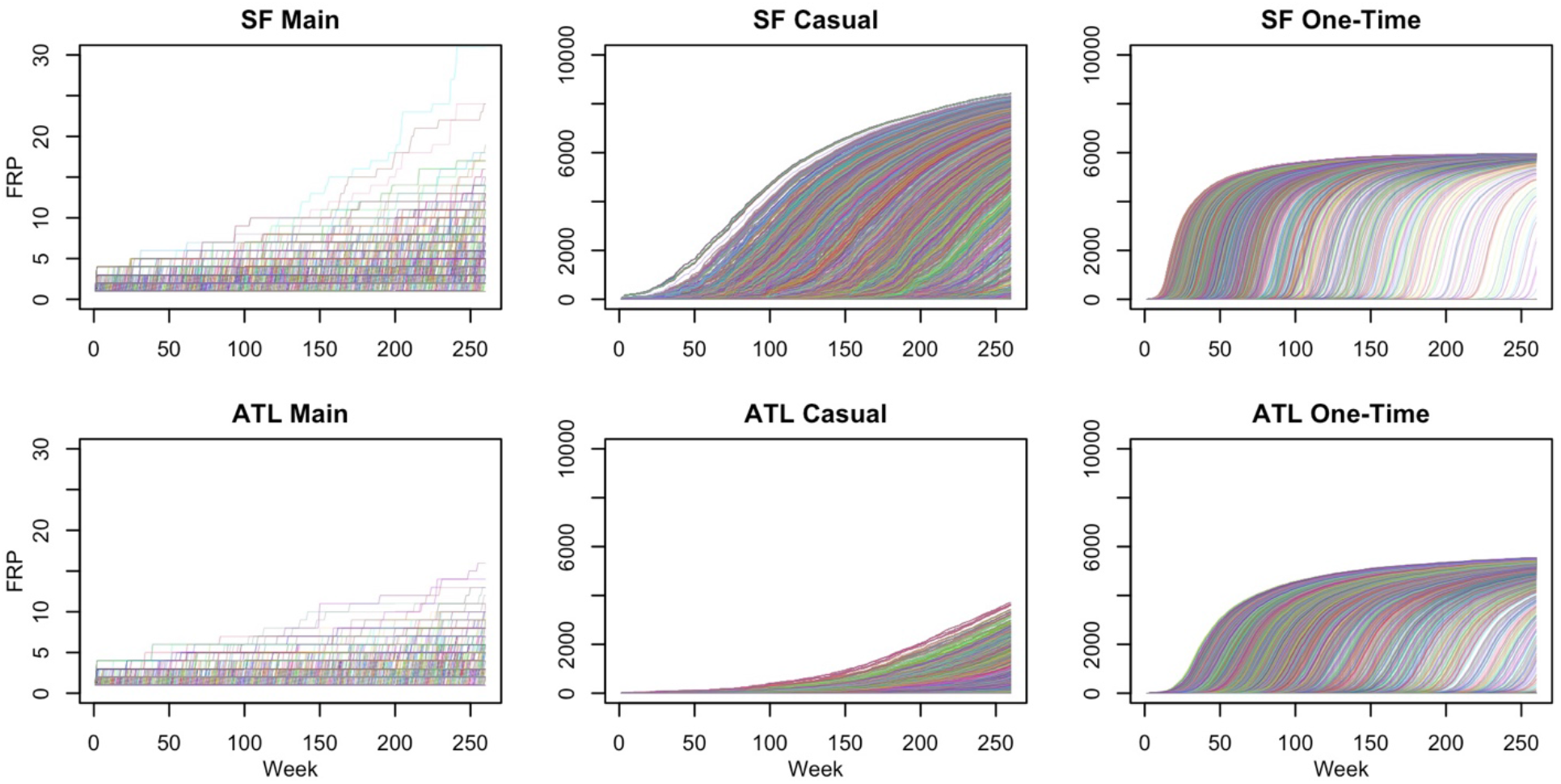
Distribution of forward reachable paths in sexual networks of MSM in San Francisco (SF) and Atlanta (ATL) by partnership type. Each line represents the trajectory of connectivity in the network over time, depending on which node started the path at time 0. Main partnerships are defined as partners considered a boyfriend, significant other, or life partner, casual as an ongoing relationship, but not a main partner, and one-time as partnerships without duration, occurring only once. The number connected, or reachable within the network, is on the Y-axis and the time step in weeks is the X-axis. The number reachable among main partnerships was limited (< 30 individuals for each city) as seen by the truncated Y-axis.

In general, for all partnership types, the FRPs reached more individuals in San Francisco than in Atlanta. Overall, within one year, on average 80% of individuals were reachable in San Francisco and 50% were reachable in Atlanta (Table 2, Column 1). Main partnership networks, characterized by long partnership durations and low degree, on average reached only a small proportion of the population in both cities (<0.02%) within one-year. Greater differences between the cities emerged in the casual and one-time partnership networks. The average percent of the population reachable after one year among casual partnerships was 1.4% and 0.08% in San Francisco and Atlanta, respectively. For one-time partnerships, the one-year average FRP was over 20% in San Francisco and about 5% in Atlanta.

The overall FRP patterns held within racial/ethnic subcategories but differed across age groups, particularly among casual partnerships. At one year, in both cities, the average FRP among casual partnerships was highest in the youngest age group (1.9% in San Francisco and 0.1% in Atlanta) and lowest in the oldest age group (1.0% in San Francisco and 0.06% in Atlanta). The five-year FRP demonstrated a similar pattern (**Figure 3**). In both cities, 15–24-year-olds had the highest FRPs and 55-64-year-olds had the lowest FRPs at every time step over the 5-years.

**Figure 3.**
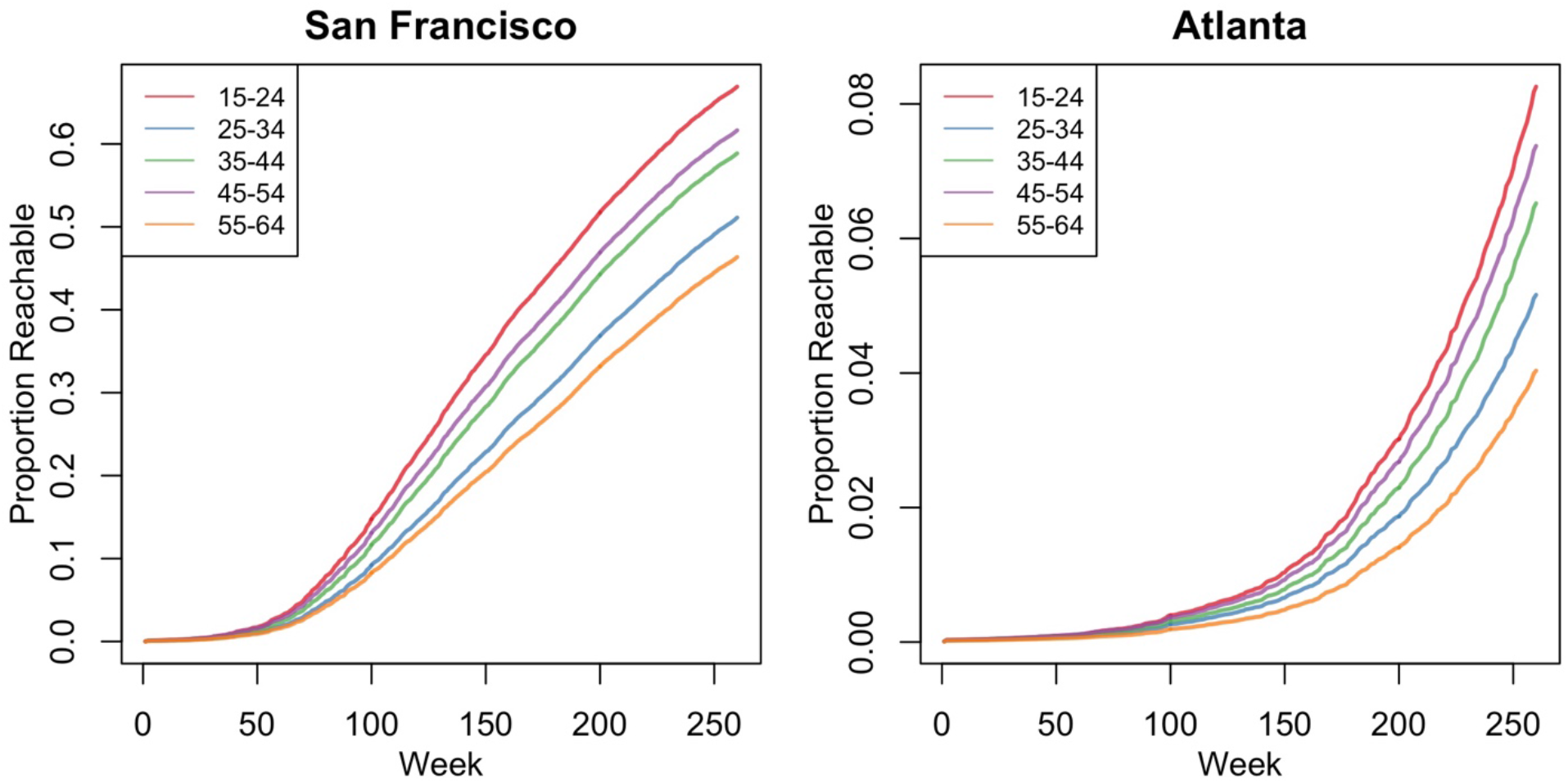
Average proportion of the San Francisco and Atlanta populations connected within sexual networks of casual partnerships of MSM over a 5-year period by age category. The Y-axis shows the average proportion of the 10,000 MSM connected, or reachable, in the network over time. The X-axis shows the time step in weeks. In San Francisco (left panel), reachability ranged from around 40-65% over 5-years, whereas in Atlanta (right panel), reachability ranged from around 4-8% over 5-years.

## DISCUSSION

In this study, we examined the FRP in sexual networks of MSM in San Francisco and Atlanta, two cities with differing HIV and STI epidemic trends. Both the mean and median FRPs were higher in San Francisco than in Atlanta, suggesting a greater epidemic potential for HIV and STIs in San Francisco. Differences in the FRP by age group emerged over time, where the oldest age group had the lowest mean FRP and the youngest age group had the highest mean FRP within casual partnerships over the five-year period. These results contrast with the cross-sectional network parameters we estimated, where the youngest age group had the lowest mean degree and the oldest age category had the highest mean degree, thus resulting in different conclusions about the epidemic potential by age depending on whether we use static or dynamic network outcomes.

Previous research on sexual networks and epidemic potential highlights that the interaction of cross-sectional network features, like degree and partnership duration, over time results in the observed temporal connectivity in a network, here measured as the FRP.^14,23^ For this study, we chose to examine the FRP in sexual networks of MSM in San Francisco and Atlanta. In San Francisco HIV rates are decreasing but STI rates are increasing faster than in Atlanta. Conversely, in Atlanta HIV rates continue to increase.^1,30,31^ Thus, we might expect these two cities to have different network structures and therefore different FRPs. Further, if the FRP does well in predicting epidemic potential, it might be used as an alternative data-driven approach to estimate epidemic potential in a network in lieu of more complex transmission modeling approaches.

Overall, the higher FRPs in San Francisco correspond to what we observe in the current bacterial STI epidemics but not the HIV epidemics between San Francisco and Atlanta. Rates of HIV have been declining in San Francisco over the last few years, a reflection of the efforts there to reduce the HIV epidemic through diagnosis and linkage to care of HIV-infected individuals and in initiating HIV-uninfected men on PrEP.^30^ In 2017, 76% of HIV-infected MSM were virally suppressed (defined as <200 copies/mL on most recent viral load test) in San Francisco.^31^ Though recent estimates were not available for Atlanta, in Georgia in 2017, 54% of MSM were estimated to be virally suppressed, suggesting better access to care in San Francisco.^31^ An undetectable viral load can effectively reduce the probability to zero of an HIV positive person transmitting to their HIV negative partner, so the more individuals on treatment the less risk of transmission within a network.^5^ San Francisco has also made great strides to increase PrEP coverage among MSM. Data from 2016 to 2018 indicate that almost all MSM were aware of PrEP, 40% used PrEP, and 35% were PrEP adherent.^30^ A 2015 study among MSM in Atlanta estimated that about 50% of the study population were aware of PrEP and 15% were expected to receive protection against HIV from PrEP usage.^37^ These advancements in HIV prevention might reduce the impact that network factors have on the risk of HIV, making the FRP less relevant for HIV. Further work is needed to understand how the interaction of network structure and public health interventions affect the epidemic potential of HIV.

Conversely to HIV, in San Francisco STI rates are on the rise and increasing faster than rates in Atlanta.^1^ The FRPs appear to reflect this dynamic better than that of HIV, with higher and faster connectivity over five years in San Francisco compared with Atlanta. It is possible that the rates of STIs in San Francisco are reflective of increased efforts to screen for STIs in addition to the HIV prevention efforts being made there. For example, the high PrEP coverage might partially explain higher STI rates through greater levels of STI screening among PrEP users. However, recent modeling work suggests that PrEP STI screening might actually decrease STI rates through better coverage of testing and treatment.^38^ Nonetheless, the individual-level network statistics and the FRPs in San Francisco indicate a greater potential for STI transmission compared to Atlanta. More research is needed to understand how the FRP correlates with different epidemic types.

For both cities, casual and one-time partnerships contributed the most to the size of the cumulative FRPs. For both these types of partnerships, the turnover rate is faster and the number of partnerships higher than with main partnerships, resulting in greater connectivity. With the assumption of 100% transmission probability, using the FRP as a model of epidemic potential means that more connectivity results in greater epidemic potential. In reality, greater connectivity does not automatically equate to greater epidemic potential. The FRP might be less relevant for infections like HIV, characterized by a lower per-act transmission probability, than for higher transmission probability STIs, especially given the centrality of one-time partnerships.^39^ Additionally, many factors likely play a role in the epidemic potential of a population.^40–42^ For example, the distribution of other risk factors in the network that were not included in this analysis might affect epidemic potential, including the distribution of the disease, who is on treatment, and PrEP use.^43–45^ A population might be very connected overall, but the disease might be concentrated within a small group that is less connected to the core of the population so that it is not as easily spread across the whole population. Nonetheless, shorter and higher numbers of partnerships and those that overlap in time do present the greatest risk for HIV and STI acquisition and transmission.^12^

Some differences in the FRP by age began to emerge within one year, especially within casual partnerships. We found that the oldest age group had the lowest mean FRP and the youngest age group had the highest mean FRP within casual partnerships at one year. These differences continued and increased over the five-year period. These results contrast with the cross-sectional network parameters we estimated, where the youngest age group had the lowest mean degree and the oldest age group had the highest mean degree. The conclusions about epidemic potential derived from these cross-sectional measures of mean degree are therefore fundamentally different than those derived from the FRP. Estimating the network over time allows for the incorporation of parameters like partnership duration, which are not accounted for in a cross-sectional network.^13,16^ The insights gained from examining the network over time are vital to accurately assessing epidemic potential, as exemplified by our results for the FRP by age.

In both cities, there were minimal differences in the FRPs between racial/ethnic categories. There could be several reasons for these results. In general, the mean degrees within each racial/ethnic category were similar to each other and also to the overall parameter estimates, though assortative mixing was more common among White/Other MSM. Therefore, we would expect the FRP to be similar between Black/Hispanic and White/Other MSM. Secondly, the small differences that there were in these parameters may not have been enough to reflect differences in network connectivity over time. Finally, because we combined racial/ethnic categories we may be missing differences between these groups that would be more apparent had we not combined them. Empirically, Black MSM tend to have more homophilous networks than non-Black MSM, but in our data Black/Hispanic respondents had lower levels of assortative mixing than White/Other respondents.^13,46,47^ This might be a reflection of combining these racial/ethnic categories, differences between our sample population and those from earlier studies, or a selection bias in our sample. Racial minorities have higher rates of HIV and STIs, which may partially be because of higher risk network structures.^46,48–50^ However, there are likely many factors playing a role in the observed disparity, like access to care^51^, which are not captured in our analysis. The FRP may not have strong explanatory power for racial disparities, similarly to how differences in sexual behaviors between racial groups have not fully explained disparities.^9,46^

### Limitations

There are several limitations to our analysis. ARTnet relied on participant-reported information, therefore, there is the potential for error in the reporting of the behavioral data we used to parameterize our network models. For instance, there might be biases in the reported number, length, and type of partnerships that favor fewer, longer term partnerships over many, shorter term ones. This could potentially result in an underestimate of the mean degrees and one-time partnership formation rates and an overestimate of the partnership durations in Table 1. Mean degree might also be underestimated because partnership data were restricted to a participant’s five most recent partnerships in the past year. If someone had many main and/or casual partners in the past year, this would not be captured in our data, but few respondents reported more than 4 partners. Another potential limitation is that we recruited a convenience sample of MSM, which might limit the generalizability of our results. In particular, online surveys may not well represent racial/ethnic minority MSM.^52^ To address this concern, we weighted our estimates in Table 1 by the race/ethnicity distribution in each city.^35^

## Conclusions

The FRP is a way to measure connectivity over time in a network. The structure of a network corresponds to differences in the FRP, as was demonstrated here in sexual networks of MSM from San Francisco and Atlanta. We found that the FRP was higher in San Francisco than in Atlanta, which might correlate to differences seen in the STI epidemics between these two cities. Future work should assess how well the FRP can predict the epidemic potential produced by an infectious disease transmission model to determine whether it is a viable alternative approach to estimating epidemic potential in sexual networks.

## Data Availability

The data is publicly available through GitHub

https://github.com/EpiModel/ARTnetData

