## Supplementary Appendix for "The HIV/STI Epidemic Potential of Dynamic Sexual Networks of Men Who Have Sex With Men in Atlanta and San Francisco"

Emeli J. Anderson,[1] Kevin M. Weiss,[1] Martina Morris,[2] Travis H. Sanchez,[1] Pragati Prasad,[1] Samuel M. Jenness[1]

[1]Department of Epidemiology, Emory University, Atlanta, GA
[2] Departments of Statistics and Sociology, University of Washington, Seattle, WA

This appendix demonstrates how the forward reachable path (FRP) measure is calculated, and how variations in network structure impact the FRP. The FRP is an individual measure that quantifies the size of the temporally ordered chain of edges (partnerships) for each node (person) in the network. For simplicity of visualization and numerical calculation, we demonstrate this with a toy network of 25 nodes using a basic network model (exponential random graph model, or ERGM) for edge formation and dissolution. The network size and model in the full paper are larger and more complex with 3 models per city, whereas here there is only one model simulated under 2 different conditions. In the Base Model we allow partnership concurrency (degree, or number of ongoing partnerships, of 2 or more). For the comparison model, we disallow concurrency. This provides a comparison between the two resulting network structures and their FRPs with the only difference between the models being concurrency.

We assume that individuals have a mean degree of 0.8 (on average 0.8 ongoing partnerships at any cross-section). This translates to an expected number of edges of 10 at any time step (edges = mean degree * N/2). On average, edges have a duration of 20 time steps. This will be a homogenous formation and dissolution model in which there is no variation in the propensity for higher activity rates or assortative mixing (in the full paper models, both are present). We simulated the network for 25 time steps, although we could have simulated the network for a longer amount of time. This would have allowed the FRP to continue to grow because it is a measure that increases monotonically over time.

**Supplemental Figure 1** below shows the FRP from the perspective of node 14 in the Base Model. Note that we could have chosen to calculate the FRP from the perspective of any node because the FRP is an individual measure. The x-axis shows the edge start time and the y-axis shows generations of hypothetical transmission events. The generation of hypothetical transmission events closely mirrors the start time of an edge, but the plot assumes that, for generation time, no more than one transmission event could occur per person per time. Supplemental Figure 1 shows all the direct and indirect network connections for node 14 across the 25 time steps. In total, node 14 has 3 direct partnerships: to nodes 11, 8, and 3. The formation of the edge with node 8 occurred later (time step 5) than nodes 3 and 11 (time step 2). Node 14 also has many indirect connections (partners of partners). For example, node 14 is one step removed from nodes 12 (through node 11) and 23 (through node 3). The FRP is the sum of both direct and indirect connections for node 14 (including node 14 itself). In this simulation, node 14 has an FRP of 13.

**Supplemental Figure 1.** The Forward Reachable Path for Node 14 in the Base Model (With Concurrency) of 25 Nodes Simulated for 25 Time Steps.

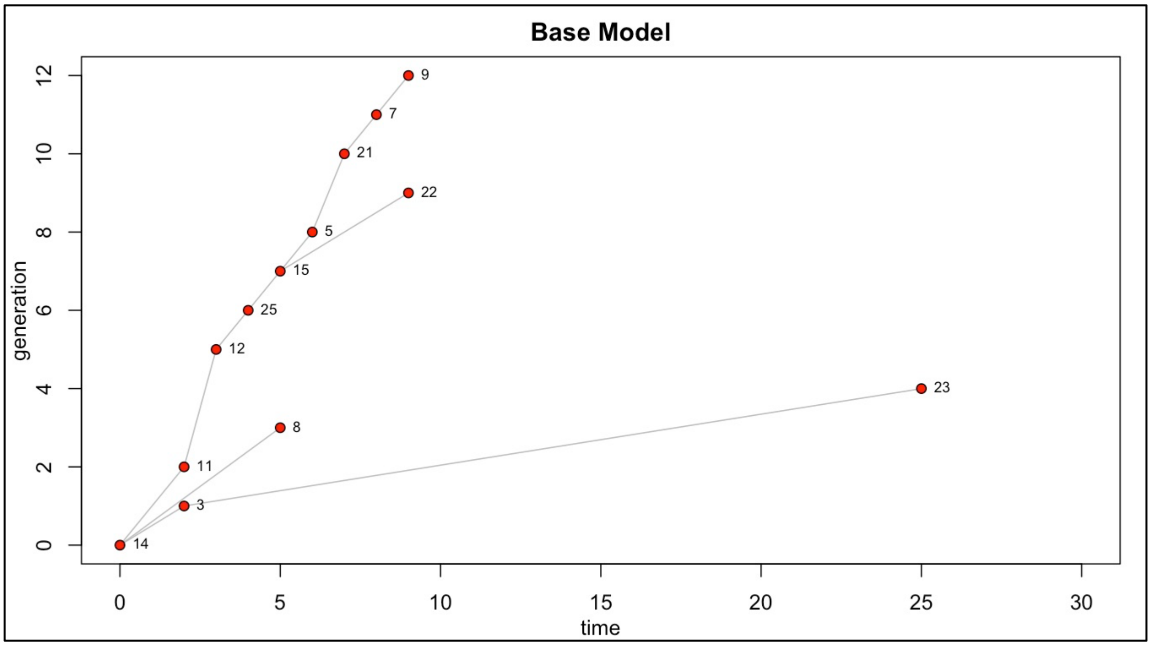


**Supplemental Figure 2** shows the underlying network that led to the FRP for node 14. It is a dynamic network, so the four panels show the network structure at times 1, 9, 17, and 25. Node 14 is highlighted in red in each panel. At time 1 (top row, left panel), node 14 is directly connected to nodes 3 and 11. Through node 11, node 14 is indirectly connected to nodes 12, 25, and so on all the way to node 9. Note that this component is connected at time 1, but node 9 is not "reached" by node 14 until time step 9 in Supplemental Figure 1. This is because there is an inherent delay of one time step as a pathogen moves from 14 to 11 to 12 to 25 and so on.

To understand the FRP in the context of this dynamic network, note the positions of nodes 22 and 23 in Supplemental Figure 1. That plot shows that they are indirect connections through nodes 15 and 3, respectively. Now locate those two nodes in each panel of Supplemental Figure 2. At time 1 (top row, left panel), nodes 22 and 23 are connected. At this point neither are in the FRP of Node 14.

By time 9 (top row, right panel), node 22 has dissolved the edge with node 23 and formed an edge with node 15, which was already indirectly connected to node 14 at time 1. Therefore, by time 9 node 22 has entered the large component surrounding node 14 and is in node 14’s FRP. Node 23 is now an isolate, not connected to any other node.

By time 17 (bottom row, left panel), the larger component has dissolved into two smaller components. At this point, nodes 14 and 22 are no longer indirectly connected, but they remain connected through the temporal FRP that was initiated earlier. Node 23 remains an isolate.

By time 25 (bottom row, right panel), node 14 has dissolved its edge with node 3, which has established a new edge with none other than node 23. Because there was an FRP already established between nodes 14 and 3, that FRP now continues on to node 23 at this late time because of this indirect connection. Note how unique and interesting this connection is between nodes 14 and 23. They were never directly or indirectly connected in a cross-sectional component at any snapshot in time. They are only connected indirectly over time. This is the essence of the FRP as a temporal measure.

**Supplemental Figure 2.** The Underlying Network that Led to the FRP for Node 14 At Time Steps 1, 9, 17, and 25.

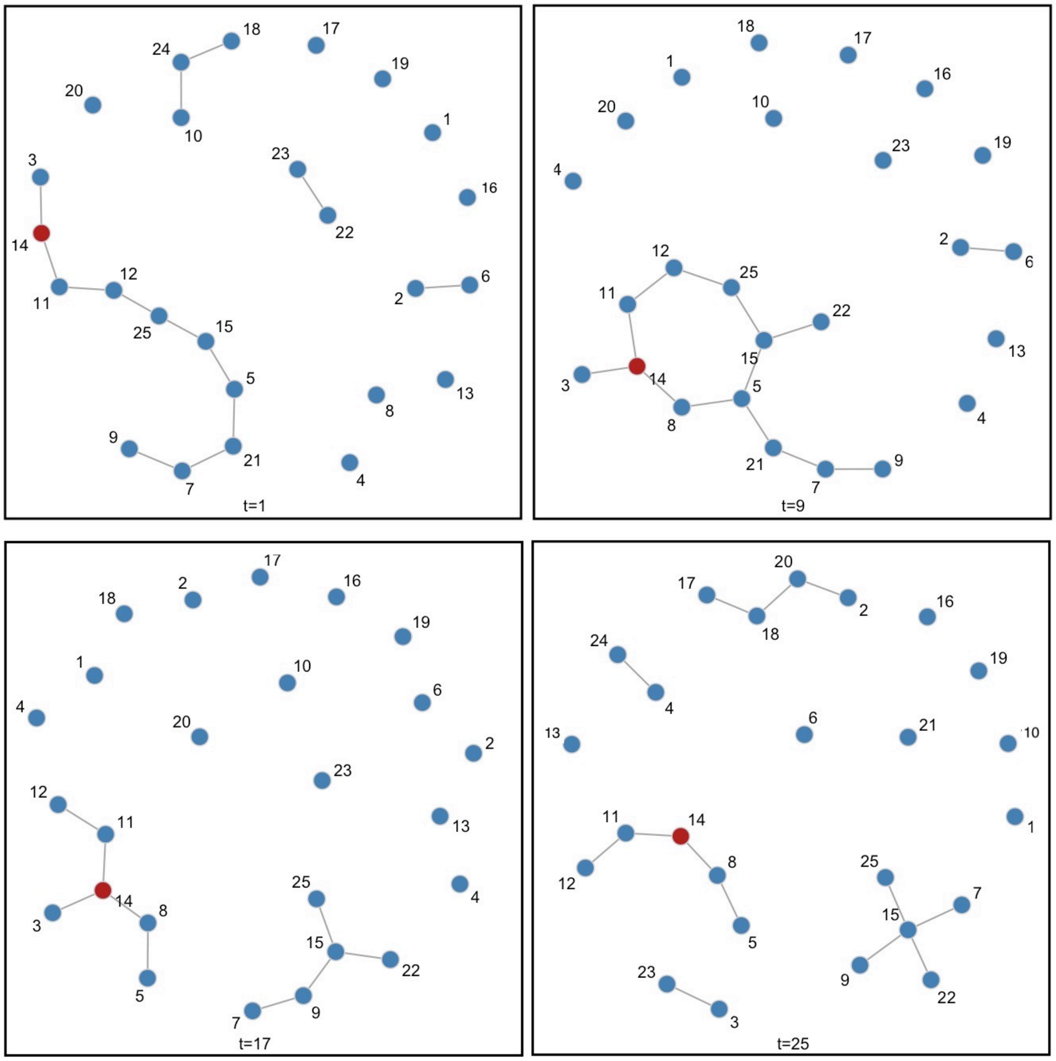

Finally, to demonstrate how changes in network structure impact the FRP in this toy example, we fit the Comparison Model in which the mean degree (and thus total expected edges) and the edge duration were the same as in the base model but concurrency (degree of 2+) was disallowed. This means that anyone in an active partnership had to end that partnership before a new one could start. The equivalent plot for the Comparison Model is shown in **Supplemental Figure 3**.

**Supplemental Figure 3.** The Forward Reachable Path for Node 14 in the Comparison Model (No Concurrency) of 25 Nodes Simulated for 25 Time Steps.

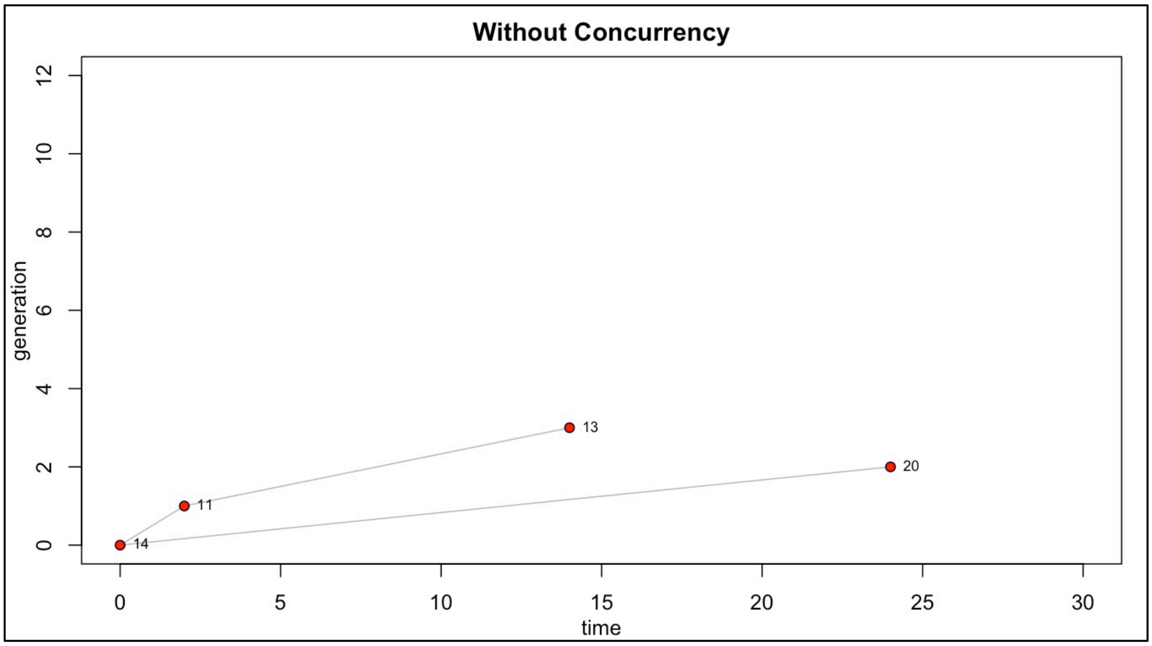


As has been demonstrated extensively in the epidemic modeling and network science literature, reductions in network degree have a substantial impact on the speed and size of epidemics. This occurs because the pathogen cannot jump across the nodes in the network with the same velocity as it can when networks are more connected through network concurrency. In this simulation, node 14 now has 2 direct partners (to nodes 11 and 20) and one indirect partnership (to node 13). The FRP for node 14 in this scenario is now 4. We hope that this demonstrates how the FRP is calculated and how changes to the network structure can impact the epidemic potential as quantified by the FRP in a simple toy model.

Code to reproduce these simulations and plots may be found at https://github.com/EpiModel/NetAnalysis-SF-ATL.
